# Boosting LLM-Assisted Diagnosis: 10-Minute LLM Tutorial Elevates Radiology Residents’ Performance in Brain MRI Interpretation

**DOI:** 10.1101/2024.07.03.24309779

**Authors:** Su Hwan Kim, Severin Schramm, Jonas Wihl, Philipp Raffler, Marlene Tahedl, Julian Canisius, Ina Luiken, Lukas Endrös, Stefan Reischl, Alexander Marka, Robert Walter, Mathias Schillmaier, Claus Zimmer, Benedikt Wiestler, Dennis M. Hedderich

## Abstract

**Purpose:** To evaluate the impact of a structured tutorial on the use of a large language model (LLM)-based search engine on radiology residents’ performance in LLM-assisted brain MRI differential diagnosis.

**Materials & Methods:** In this retrospective study, nine radiology residents determined the three most likely differential diagnoses for three sets of ten brain MRI cases with a challenging yet definite diagnosis. Each set of cases was assessed 1) with the support of conventional internet search, 2) using an LLM-based search engine (© Perplexity AI) without prior training, or 3) with LLM assistance after a structured 10-minute tutorial on how to effectively use the tool for differential diagnosis. The tutorial content was based on the results of two studies on LLM-assisted radiological diagnosis and included a prompt template. Reader responses were rated using a binary and numeric scoring system. Reading times were tracked and confidence levels were recorded on a 5-point Likert scale. Binary and numeric scores were analyzed using chi-square tests and pairwise Mann-Whitney U tests each. Search engine logs were examined to quantify user interaction metrics, and to identify hallucinations and misinterpretations in LLM responses.

**Results:** Radiology residents achieved the highest accuracy when employing the LLM-based search engine following the tutorial, indicating the correct diagnosis among the top three differential diagnoses in 62.5% of cases (55/88). This was followed by the LLM-assisted workflow before the tutorial (44.8%; 39/87) and the conventional internet search workflow (32.2%; 28/87). The LLM tutorial led to significantly higher performance (binary scores: p = 0.042, numeric scores: p = 0.016) and confidence (p = 0.006) but resulted in no relevant differences in reading times. Hallucinations were found in 5.1% of LLM queries.

**Conclusion:** A structured 10-minute LLM tutorial increased performance and confidence levels in LLM-assisted brain MRI differential diagnosis among radiology residents.

**Clinical Relevance Statement:** Our findings highlight the considerable benefits that even low-cost, low-effort educational interventions on LLMs can provide. Integrating LLM education in radiology training programs could augment practitioners’ capacity to harness AI technologies effectively.

## Introduction

Large language models (LLMs) are advanced artificial intelligence (AI) systems capable of processing and generating human language. Trained on vast amounts of text data and based on an innovative transformer architecture, these models have demonstrated remarkable performance in various tasks across sectors (1).

With the rapid technological advancements of LLMs in recent years, numerous studies have explored applications of LLMs in radiological workflows. These include the definition of imaging protocols (2–4), performing differential diagnosis based on case presentations (5–9), error checking in radiology reports (10), generation of impressions in radiology reports (11,12), information extraction from free-text radiology reports (13–15) and more. Yet, despite the promising applications, the integration and adoption of LLMs in radiology is not without challenges. Data privacy concerns, bias and error propagation, lack of contextual understanding, and overreliance have been pointed out as relevant limitations of LLMs (1,16–19). Against this background, the critical role of educating healthcare professionals on the appropriate use and potential pitfalls of LLMs has been emphasized (20–22). This may include training in prompt engineering, which describes the strategic crafting of a textual instruction that serves as input to LLMs (22).

One area where insufficient human oversight of LLMs could lead to clinical errors is radiological differential diagnosis. An earlier study on human-LLM collaboration in brain MRI differential diagnosis found that inadequate formulation of prompts can result in misleading LLM outputs, and lacking critical validation of LLM responses can lead to incorrect conclusions (23).

However, whether and how radiology readers can be trained to more effectively apply LLMs in radiological differential diagnosis has not been investigated yet. This study therefore aimed to evaluate the impact of a structured LLM tutorial on the performance of radiology residents in brain MRI differential diagnosis.

## Methods

Informed patient consent was waived by the Ethics Committee of the Technical University of Munich.

### Study Sample

Thirty challenging brain MRI exams acquired between 01/01/2016 and 12/31/2023 were selected from the local Picture Archiving and Communication System (PACS) system and randomized into three sets (Figure 1). Included exams were deemed as sufficiently complex for use in radiological board certification exams by two board-certified neuroradiologists (DMH and BW) and contained an abnormal finding with a confirmed diagnosis (histopathologically or through independent agreement of at least two neuroradiologists). In each exam, one or more arrows marked the image finding in question. The included exams have been published previously (24). A case overview is provided in Supplement 1.

**Figure 1:**
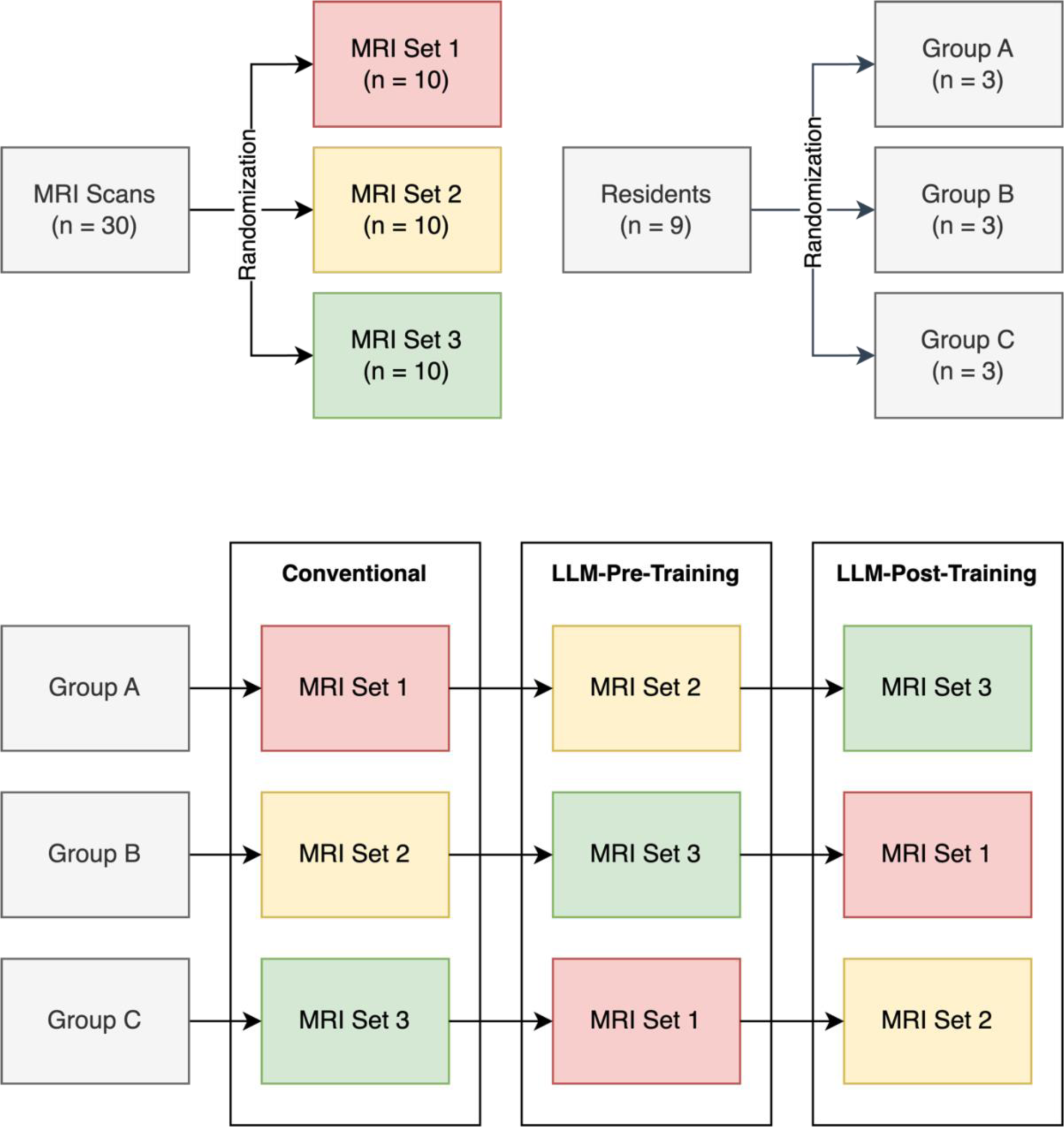
Study Design.

Nine radiology residents with less than six months of neuroradiology experience were recruited from the local departments of radiology and neuroradiology and randomized into three groups (Table 1). Informed consent was provided by all participants.

**Table 1:** Overview of readers.

| Trait | Value |
| --- | --- |
| Total Number of Readers | 9 |
| Gender Distribution | 7 Male (77.8%) / 2 Female (22.2%) |
| Mean radiology experience (in months) | 18.56 ± 15.93 |
| Mean neuroradiology experience (in months) | 2.22 ± 2.05 |
| Readers who have used LLMs before | 6/9 (66.7%) |
| Readers who have used LLMs for diagnosis before | 1/9 (11.1%) |

### Study Design

Over the course of three sessions, each reader assessed three sets of ten brain MRI cases with varying workflows and provided up to three differential diagnoses for the annotated image findings, ranked by likelihood. Each case was reviewed only once per reader. To control for the confounding effects of case difficulty, each set of cases was assessed by the same number of readers with each workflow (Figure 1). For every case, demographic and condensed medical history was provided. Sessions took place between 01/03/2024 and 22/05/2024.

First, conventional internet research was conducted to support differential diagnosis, either using web-based search engines, e.g. Google Search, or directly accessing trusted websites (Conventional). Residents were instructed to behave as they would in a clinical routine setting, mimicking the current practice in clinical care. Second, readers utilized an LLM-based search engine (© Perplexity AI Inc., San Francisco, USA) but didn’t receive any training beforehand (LLM-Pre-Training). PerplexityAI had been chosen as LLM interface for its ability to access real-time web content and provide source citations. Search queries were powered by GPT-4-Turbo (Generative Pre-trained Transformer 4 Turbo) by OpenAI. Third, another subset of ten cases was evaluated with the assistance of PerplexityAI. This time, however, the session was preceded by a structured tutorial on how to effectively use the tool (LLM-Post-Training). Tutorial details are provided below. In both LLM-assisted workflows, participants were allowed to conduct additional internet search to validate LLM suggestions.

Reading times were recorded using a time tracking software (Toggl Track, © Toggl OÜ, Talinn, Estonia). Confidence levels were documented for each case on a 5-point Likert scale (1: not at all confident, 5: very confident). Following the second and third session, readers completed questionnaires to evaluate the experience with the LLM-assisted workflow.

### LLM Tutorial

> In a short tutorial of no more than 10 minutes, readers were given tips on how to effectively utilize the LLM-based search engine. The content of the tutorial was based on two earlier studies on the application of LLMs for brain MRI differential diagnosis. One study evaluated the contribution of varying multimodal input elements on the diagnostic performance of GPT4(V) and identified the textual description of radiological image findings as the key element (24). The other demonstrated superior accuracy of LLM-assisted differential diagnosis over a workflow supported by a conventional search engine, but also determined several pitfalls in human-LLM interaction (23). The full script of the tutorial is provided below:

> “A detailed description of image findings is by far the most important factor for accurate LLM responses. The description should include details about location, contrast enhancement, morphology, size and more. An accurate description of the finding location is particularly critical, an inaccurate specification of the location can result in misleading suggestions.

> Providing relevant information about the medical history can improve the accuracy of LLM responses. However, clinical information unrelated to the image finding might result in misleading LLM outputs. Therefore only clinical information deemed to be relevant for the image finding in question should be provided.

> Uploading screenshots of key image findings can help improve LLM responses, although their effect is only marginal.

> Use of connotative terminology can lead to bias and should be avoided (e.g. the term ‘juxtacortical’ is strongly associated with multiple sclerosis).

> Instructions can be made regarding the extent (number of differential diagnoses mentioned) and format (bullet points, table) of the LLM output.“

Readers were further encouraged to use the following prompt template:

> “You are a senior neuroradiologist. Below, you will find information regarding a brain MRI scan. Based on this information, identify the three most likely differential diagnoses, ranked by their likelihood. Present your findings in a table format with the following columns: ‘Rank’, ‘Differential Diagnosis’, and ‘Explanation’.

> [Medical history] [Image description]”

### Analysis

To ensure performance did not solely depend on prior knowledge but the quality of research, cases where the correct diagnosis could be determined confidently without further research were excluded.

Accuracy of differential diagnoses was determined using two different scoring systems, as described previously (23). The first method used a binary scoring system, where responses were labeled as “correct” if the correct diagnosis was included among the submitted differentials, and “incorrect” if it was not. The second approach assigned scores ranging from 0 to 3 based on the rank of the correct diagnosis within the response (0: correct diagnosis not included, 1: correct diagnosis ranked third, 2: correct diagnosis ranked second, 3: correct diagnosis ranked first). Cases where a correct but less granular response was indicated were rated in consensus (by SHK and SS). For binary scores, a chi-square test was initially applied across all groups, followed by pairwise chi-square tests. For numeric scores and confidence levels, a Kruskal-Wallis test was used to assess differences among all groups, with subsequent pairwise comparisons conducted using the Mann-Whitney U test. To control for false discovery rates, p-values were adjusted using the Benjamini-Hochberg procedure for both scores and confidence. Reading times were analyzed using an ANOVA test across all workflows, followed by pairwise t-tests. The significance level was set at p < 0.05. 5-point Likert-scale questionnaire results are reported using descriptive statistics.

Logs of the LLM-based search engine (Perplexity AI) were examined to quantify the number of queries and source references. Queries were categorized by query type (keyword-based vs instruction-based) and by content (differential diagnosis, radiographic features, sample images, anatomy, other). Sources were classified into journal articles and other online sources. The content of LLM responses were screened for incorrect or inconsistent information by two radiology residents (SHK and SS; 1.5 and 2.5 years of experience in reading brain MRI exams) and confirmed by a certified neuroradiologist (DH). As described previously (25), incorrect responses were classified into hallucinations (inconsistent with widely accepted radiological knowledge), misinterpretations (miscomprehending a question and giving contextually irrelevant replies), and clarifications (lacking comprehension of a prompt requiring its rephrasing). Radiographic features described in LLM responses were checked against reference articles of www.radiopaedia.org, which is a validated source of radiological knowledge.

Data curation, analysis and visualization were performed using Python (version 3.9.7).

## Results

8 out of 270 responses (3.0%) were excluded from the analysis as readers were able to determine the correct diagnosis confidently without requiring further research. A sample LLM query and its results are shown in Figure 2. 12 out of 262 cases (4.6%) required a consensus decision because the reader provided a correct but less specific diagnosis (e.g. “encephalitis” was counted as correct in a case of limbic encephalitis).

**Figure 2:**
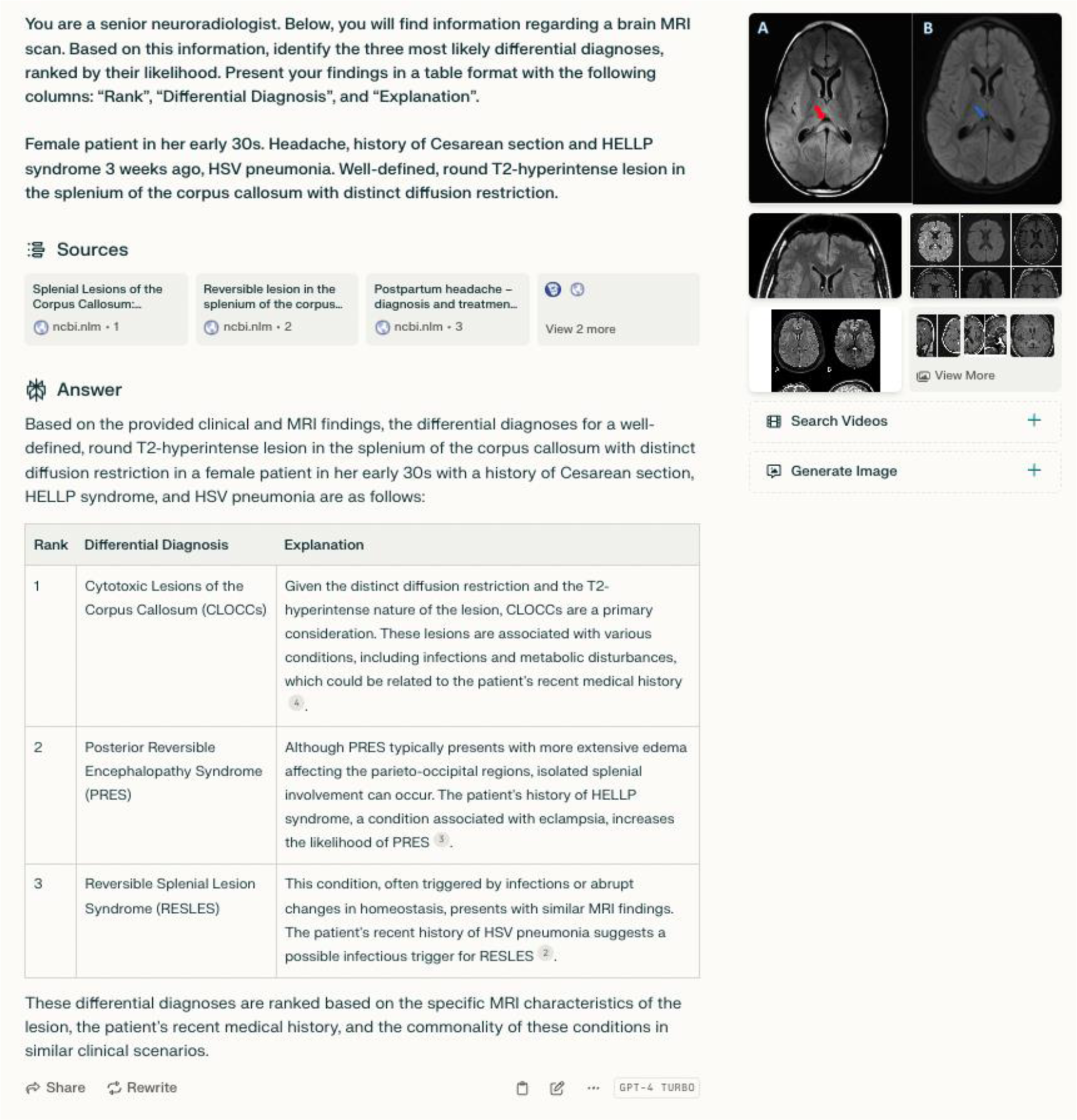
Screenshot of the PerplexityAI user interface. The correct diagnosis in this case was cytotoxic lesion of the corpus callosum (CLOCC).

### Binary and Numeric Scores

Based on the binary scoring system, 62.5% (55/88) of responses in the LLM-Post-Training workflow were correct, compared to 44.8% (39/87) in the LLM-Pre-Training and 32.2% (28/87) in the Conventional group (Figure 3). An initial chi-square test across all groups indicated a significant overall difference (p < 0.001). Subsequent pairwise chi-square tests showed significant differences between LLM-Pre-Training and LLM-Post-Training (p = 0.042), as well as between LLM-Post-Training and Conventional (p < 0.001), but not between LLM-Pre-Training and Conventional (p = 0.119) (Table 2).

**Figure 3:**
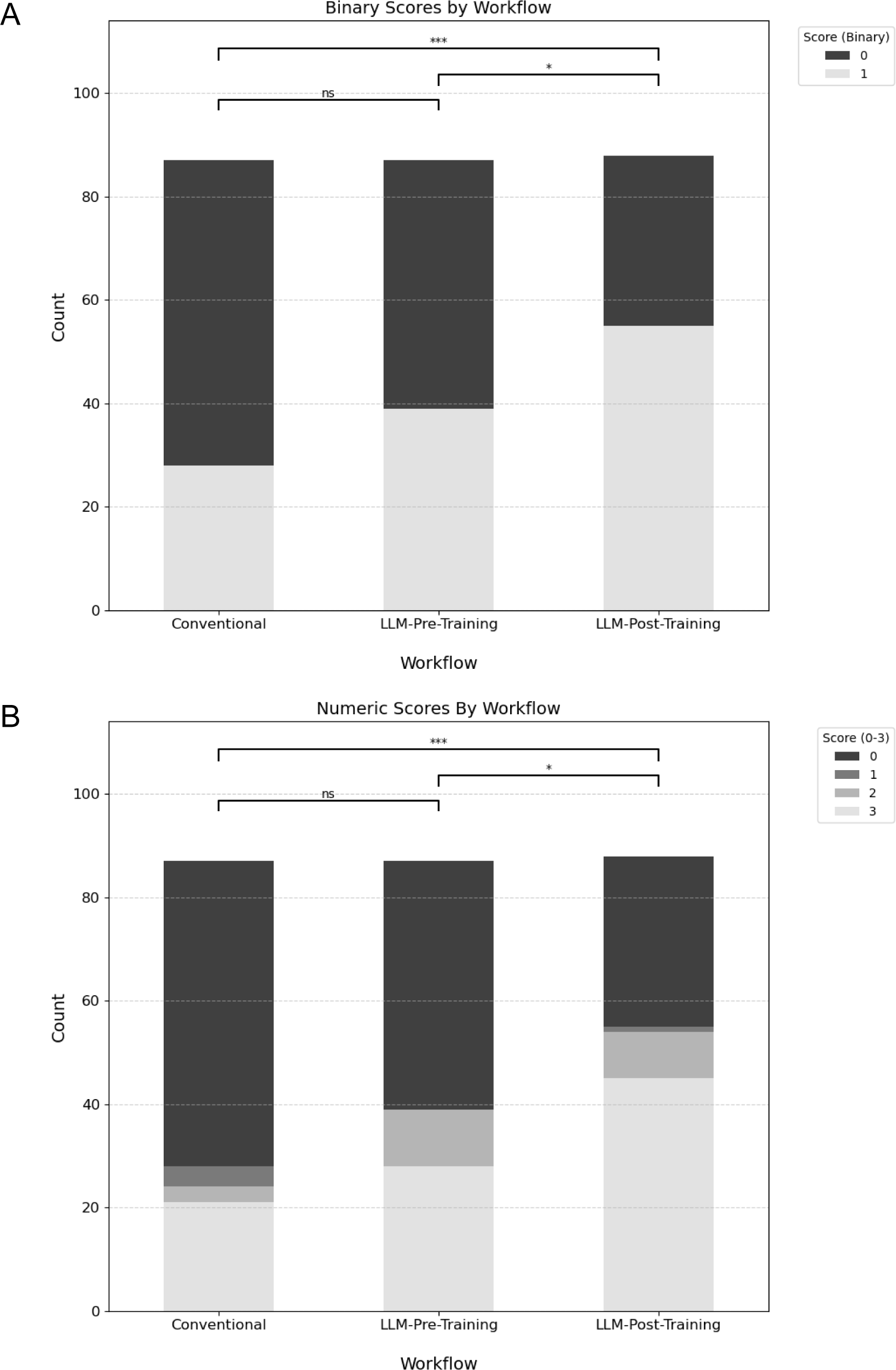
Diagnostic performance by workflow. A: Binary scores. Responses were classified as either correct (1) or incorrect (0). B: Numeric scores. Responses were assigned a score between 0 and 3, depending on the rank of the correct diagnosis within the response (3: correct diagnosis ranked first, 0: correct diagnosis not included in response). * p < 0.05. *** p < 0.001. ns: not significant.

**Table 2:** Pairwise testing for inter-group differences in binary scores. Adjusted p-values have been corrected for a false-discovery rate of 0.05.

| Comparison | Chi2 Statistic | p | p (adjusted) |
| --- | --- | --- | --- |
| LLM-Pre-Training vs LLM-Post-Training | 4.81 | 0.028 | **0.042** |
| LLM-Pre-Training vs Conventional | 2.43 | 0.119 | 0.119 |
| LLM-Post-Training vs Conventional | 14.93 | < 0.001 | **< 0.001** |

Comparison of numeric scores revealed a median score of 3 in the LLM-Post-Training group, compared to 0 for both the LLM-Pre-Training and Conventional groups (Figure 3). The Kruskal-Wallis test confirmed significant overall differences among the workflows (p < 0.001). Pairwise comparisons using the Mann-Whitney U test revealed significant differences between LLM-Pre-Training and LLM-Post-Training (p = 0.016) and between LLM-Post-Training and Conventional (p < 0.001), but not between LLM-Pre-Training and Conventional (p = 0.092) (Table 3).

**Table 3:**
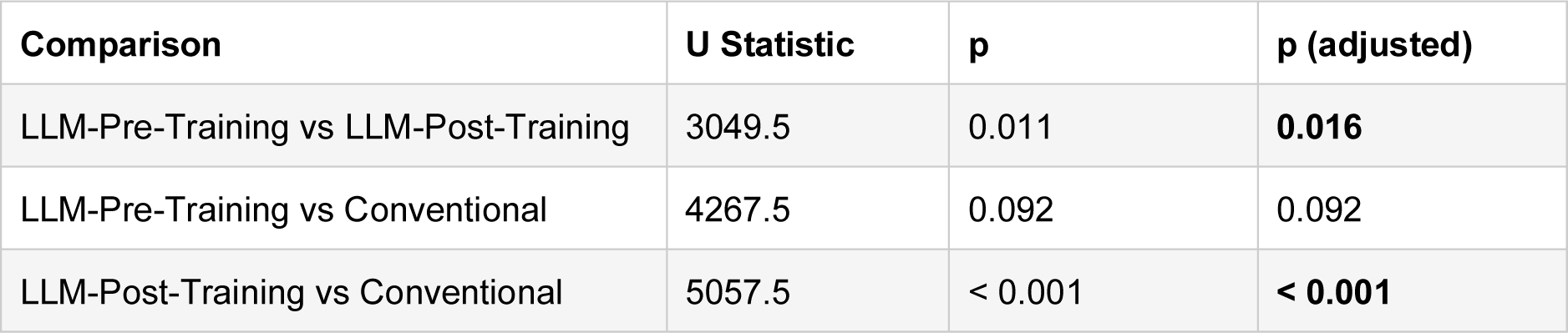
Pairwise testing for inter-group differences in numeric scores. Adjusted p-values have been corrected for a false-discovery rate of 0.05.

| Comparison | U Statistic | p | p (adjusted) |
| --- | --- | --- | --- |
| LLM-Pre-Training vs LLM-Post-Training | 3049.5 | 0.011 | **0.016** |
| LLM-Pre-Training vs Conventional | 4267.5 | 0.092 | 0.092 |
| LLM-Post-Training vs Conventional | 5057.5 | < 0.001 | **< 0.001** |

### Confidence

Median confidence ratings were highest in LLM-Post-Training (median = 4), followed by LLM-Pre-Training (median = 3) and Conventional (median = 3). The proportion of high or very high confidence ratings (4 or 5) was 18% for Conventional, 31% for LLM-Pre-Training, and 54% for LLM-Post-Training (Figure 4). The Kruskal-Wallis test showed a significant overall difference in confidence (H = 27.20, p < 0.001). Pairwise Mann-Whitney U tests revealed a highly significant difference between Conventional and LLM-Pre-Training (p = 0.006), LLM-Pre-Training and LLM-Post-Training (p = 0.006) as well as between Conventional and LLM-Post-Training (p < 0.001).

**Figure 4:**
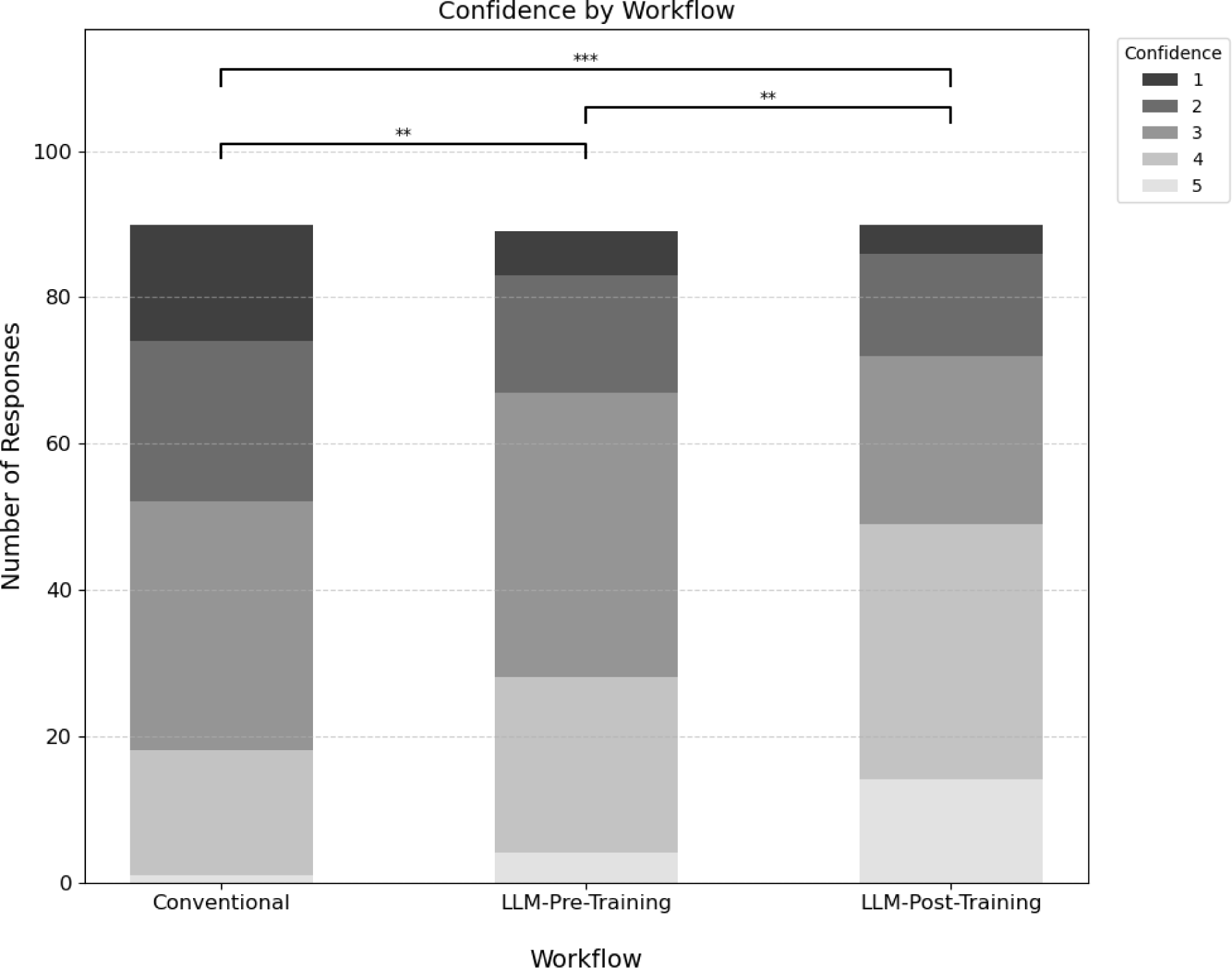
Confidence (5-point Likert scale rating) by workflow. ** p < 0.01. *** p < 0.001.

### Reading Times

Mean reading times amounted to 07:43 min (Conventional), 08:59 min (LLM-Pre-Training) and 08:35 min (LLM-Post-Training) (Figure 5). An ANOVA test showed a statistically significant overall difference (p = 0.030). Pairwise t-tests showed significant differences between Conventional and LLM-Pre-Training (p = 0.013), but not between LLM-Pre-Training and LLM-Post-Training (p = 0.403) or between Conventional and LLM-Post-Training (p = 0.069).

**Figure 5:**
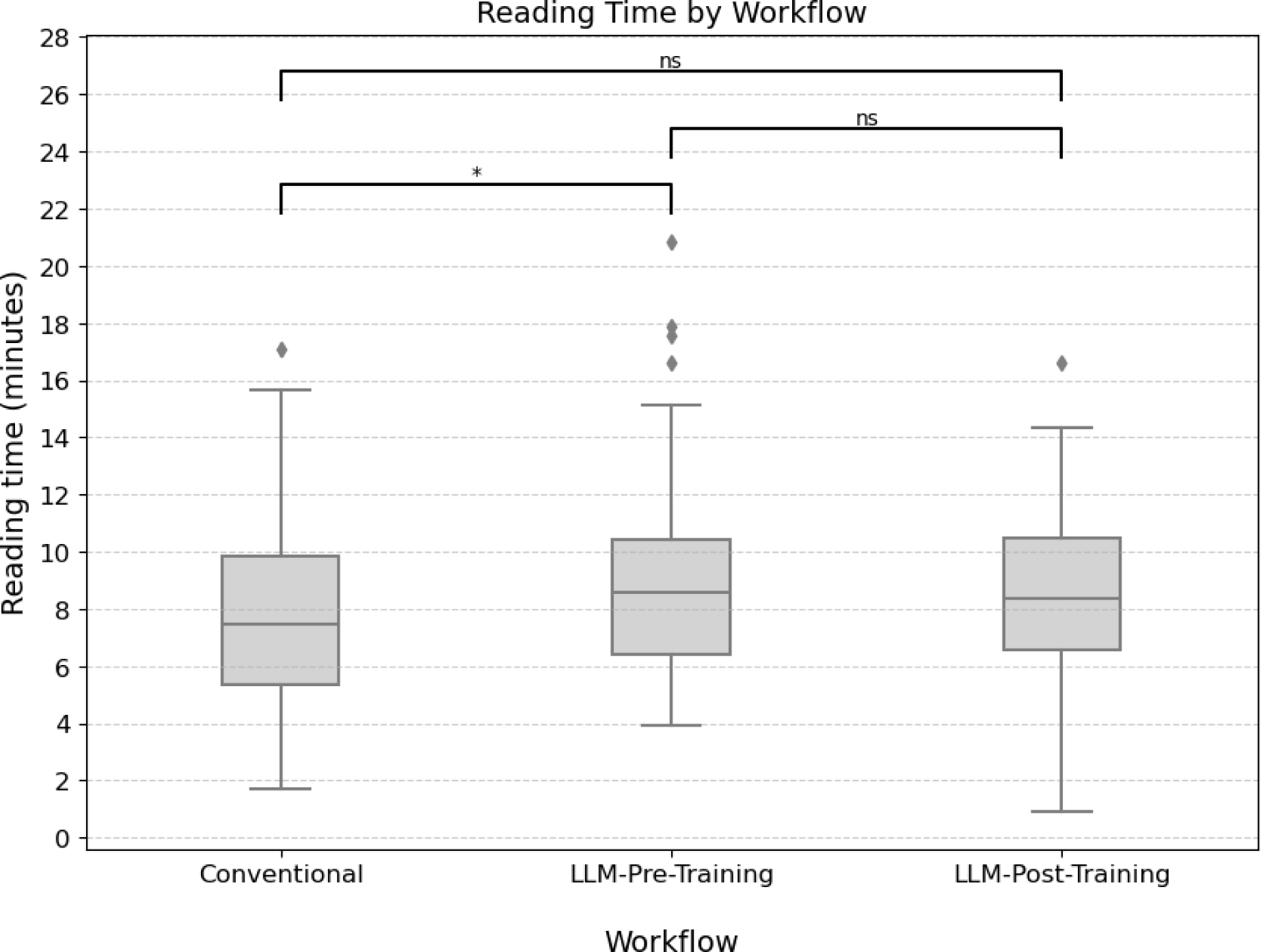
Reading time by workflow. * p < 0.05. ns: not significant.

**Figure 6:**
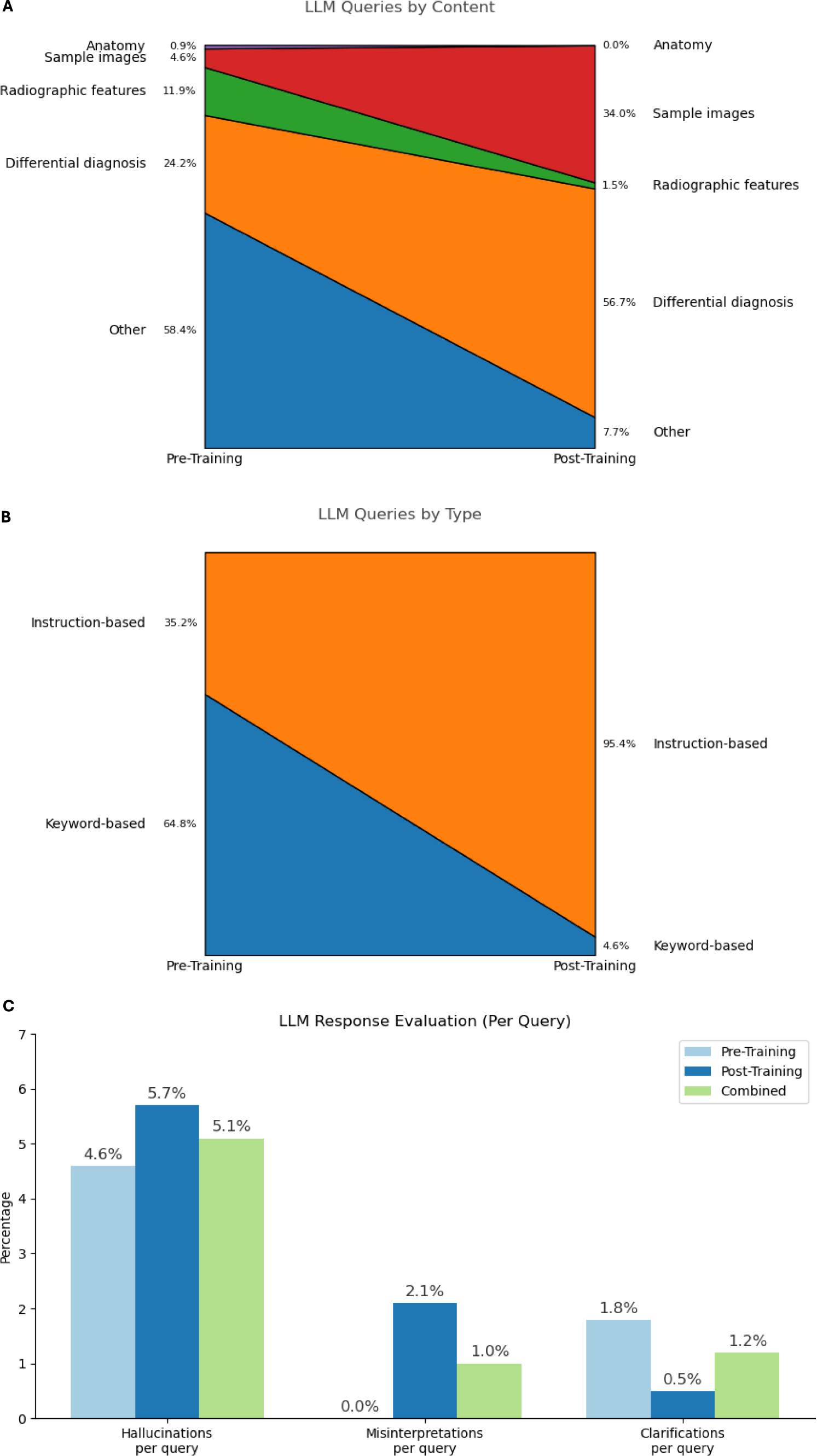
Evaluation of LLM queries and responses. A: LLM queries by content. B: LLM queries by type. C: Relative frequency of hallucinations, misinterpretations and clarifications per query.

### Questionnaire Results

The proportion of readers satisfied or very satisfied with the LLM-assisted diagnostic workflow increased from 55.6% (5/9) to 88.9% (8/9) following the tutorial. 55.6% (5/9) of readers indicated they would consider using the tool in clinical practice before the training, compared to 88.9% (8/9) after the training. 88.9% (8/9) of readers found the tutorial helpful or very helpful.

### Reader Feedback and Observations

Before the tutorial, most readers tended to use keyword-based queries rather than detailed instructions, similar to how conventional search engines operate. In general, the LLM tool was found particularly useful in generating an initial list of possible differential diagnoses to be evaluated through additional searches. In addition, the possibility to pose follow-up questions to initial query results was perceived as an advantage over conventional search engines. When using the provided prompt template, readers overwhelmingly appreciated the concise tabular format of the results which also included the rationale for the suggestion. However, some readers struggled to formulate accurate image descriptions, owing to their insufficient knowledge of neuroanatomy and brain MRI sequences.

### LLM Response Evaluation

A total of 413 LLM queries in 169 patient cases were examined (2.44 queries per case). In 11 out of 180 cases, LLM logs were not available because the user did not perform any LLM queries or because logs could not be retrieved due to technical errors of PerplexityAI. 5.8% of queries included incorrect inputs, such as inaccurate descriptions of finding locations or imaging characteristics. 7.3% of queries included screenshots of MRI findings.

The proportion of instruction-based queries increased substantially from 35.2% to 95.4% after the tutorial, while keyword queries decreased inversely from 64.8% to 4.6%. Whereas the majority of queries prior to the training were classified as “Other” (58.4%), most queries following the tutorial were directed at relevant differential diagnoses (56.7%) and sample images of those (34.0%). Overall, 45.3% of sources indicated by PerplexityAI were peer-reviewed journal articles, with only minor differences between queries before and after the tutorial (49.2% and 40.6% each).

Hallucinations were observed in 5.1% of LLM queries (12.4% of cases; 20 responses in total). The LLM tutorial resulted in only a minor change in hallucination frequency (Pre-Training: 4.6%, Post-Training: 5.7%). 35.0% of hallucinations involved the misinterpretation of MRI screenshots provided as input or the LLM returning sample MRI images irrelevant to the context of the query. Hallucination details are provided in Supplement 2. Misinterpretations were found in 1.0% of queries (2.4% of cases), while clarifications occurred in 1.2% of queries (3.0% of cases).

## Discussion

This study evaluated the impact of a structured 10-minute LLM tutorial on the performance of radiology residents in LLM-assisted brain MRI differential diagnosis. We found that readers displayed higher performance, confidence levels and overall satisfaction after completing the tutorial. Compared to differential diagnosis supported by conventional internet search, both LLM-assisted workflows resulted in better performance, although only the post-training workflow showed a statistically significant difference.

Analysis of reader-LLM interactions revealed that following the tutorial, almost all queries were phrased as specific instructions, whereas most queries before the training consisted of mere keywords, resembling conventional search engine queries. This observation is consistent with “Jakob’s Law” which is a well-known phenomenon in user experience (UX) stating that users prefer systems to behave like other familiar ones (26). Similar to hallucination rates described previously (25), we found statements inconsistent with widely accepted medical knowledge in 5.1% of LLM responses. Many of these involved incorrect interpretations of MRI screenshots provided as input, confirming earlier studies demonstrating low performance of current state-of-the art LLMs in diagnostic tasks based on radiological images (27–30). Interestingly, hallucinations were even found with PerplexityAI, which – unlike other Chatbots such as ChatGPT - combines LLMs with real-time information retrieval from the internet to support its answers with relevant sources (31). Additional research is needed to develop further safeguarding measures against potentially harmful effects of hallucinations in clinical LLM applications. Notably, in 5.8% of queries misleading responses were generated not because of undesired LLM behavior, but due to incorrect finding descriptions provided by readers, emphasizing the essential role of conventional radiological skills in effectively employing LLMs for diagnostic tasks.

Our findings indicate that even minor, low-cost educational interventions for LLMs can yield remarkable outcomes, and support the notion that courses focused on the practical application of AI should become a core part of medical curricula and training programs (32–34). Yet, given the novelty of the technology, validated educational content on the effective utilization of LLMs for specific clinical tasks is extremely scarce. The tutorial provided to the readers in this work was based on two previous studies specifically focusing on human-LLM collaboration and prompt engineering in brain MRI differential diagnosis (23,24). As the corpus of scientific evidence on LLM applications grows, medical societies should provide guidelines and courses on their appropriate use. Furthermore, platforms should be created to allow for healthcare professionals to exchange validated and effective prompts, similar to previous initiatives by radiological societies for sharing structured reporting templates (35–37).

Unlike diagnostic performance, reading times did not improve with either of the LLM-assisted workflows. As prior work on AI-based image analysis algorithms and structured reporting has illustrated (38,39), integration of technologies into the local IT infrastructure is critical for user acceptance and can boost efficiency. Vendors of radiology reporting solutions and Picture Archiving and Communication Systems (PACS) should explore ways to seamlessly embed LLM-based features supporting differential diagnosis and other tasks.

### Limitations

The following limitations need to be acknowledged.

First, only radiology residents with very little neuroradiology experience were included. This study design ensured that most cases could not be solved by readers with prior knowledge alone and allowed to investigate the isolated effect of distinct web research workflows, but limits generalizability of the findings to more experienced readers. Yet, our observation that inexperienced readers showed performance improvements ensuing the LLM tutorial despite struggling to formulate image descriptions suggests that moderately experienced readers, who are proficient enough to create accurate finding descriptions but not yet skilled enough to conduct differential diagnoses without assistance, might benefit even more.

Second, the findings in question were presented with annotations to isolate readers’ classification performance, but this approach reduced the realism of the scenario. It is likely that in actual practice, some of the more subtle findings would have been missed, as remarked by several readers.

Third, this study employed only a single LLM (GPT-4 by OpenAI) accessed through a specialized search engine (PerplexityAI). Future studies should compare several closed-source and open-source LLMs with respect to their utility in supporting radiology readers in differential diagnosis, including ones fine-tuned with domain-specific training data.

In conclusion, a concise but structured 10-minute LLM tutorial increased performance and confidence levels in LLM-assisted brain MRI differential diagnosis among radiology residents. These findings highlight the considerable benefits that even low-cost, low-effort educational interventions on LLMs can provide.

## Data Availability

All data produced in the present study are available upon reasonable request to the authors

---

**Supplement 1:**
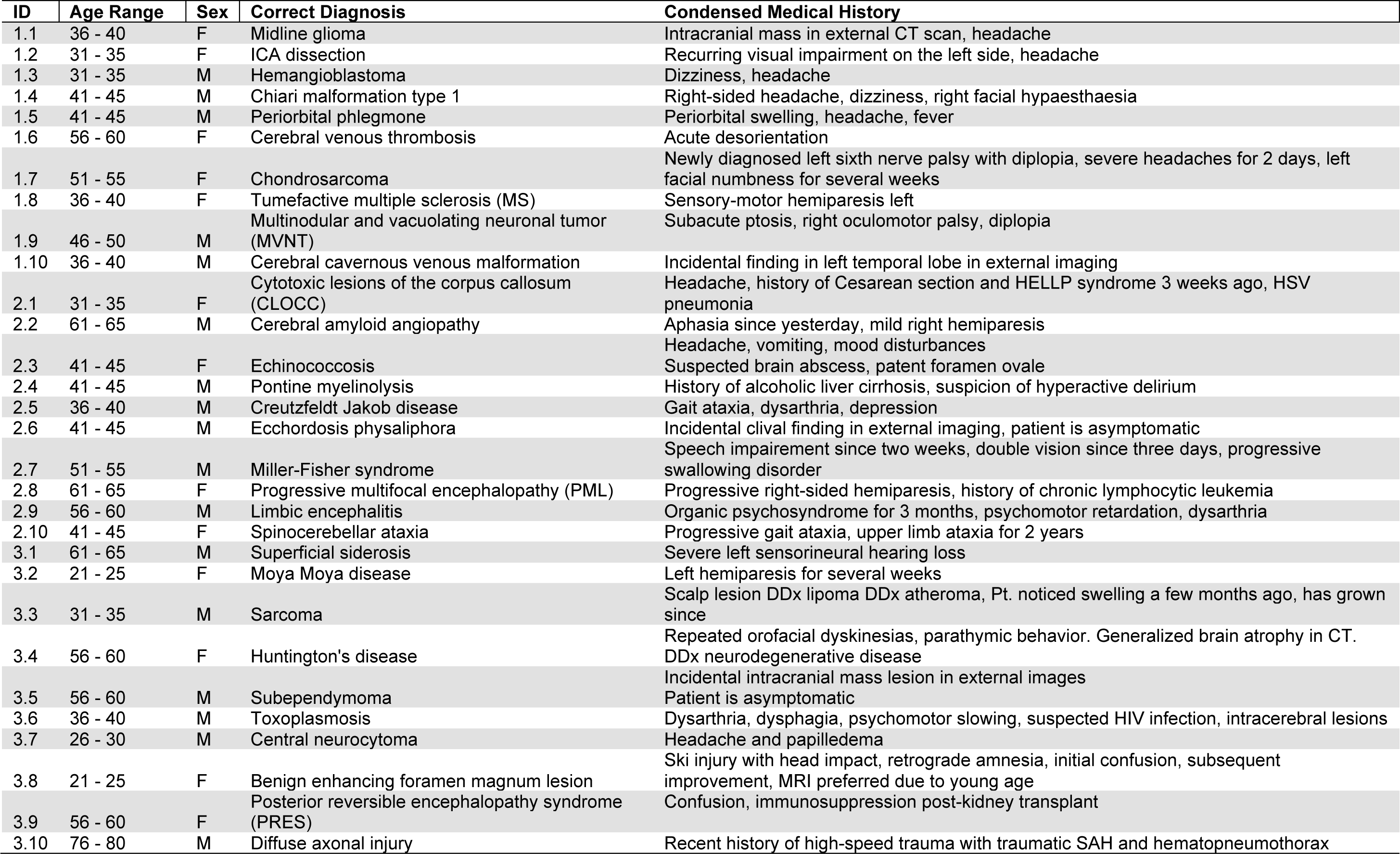
Case Overview.

**Supplement 2:**
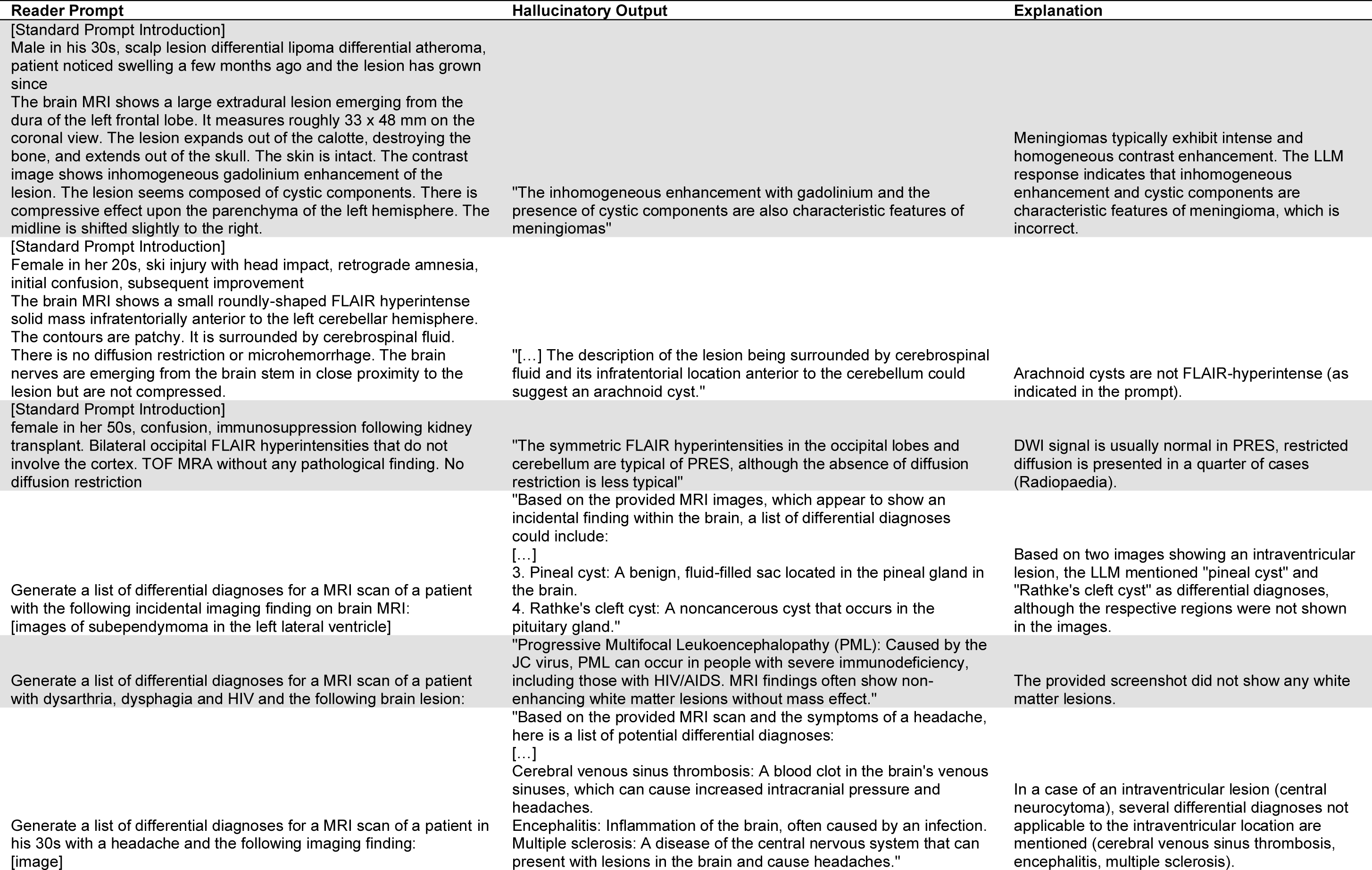
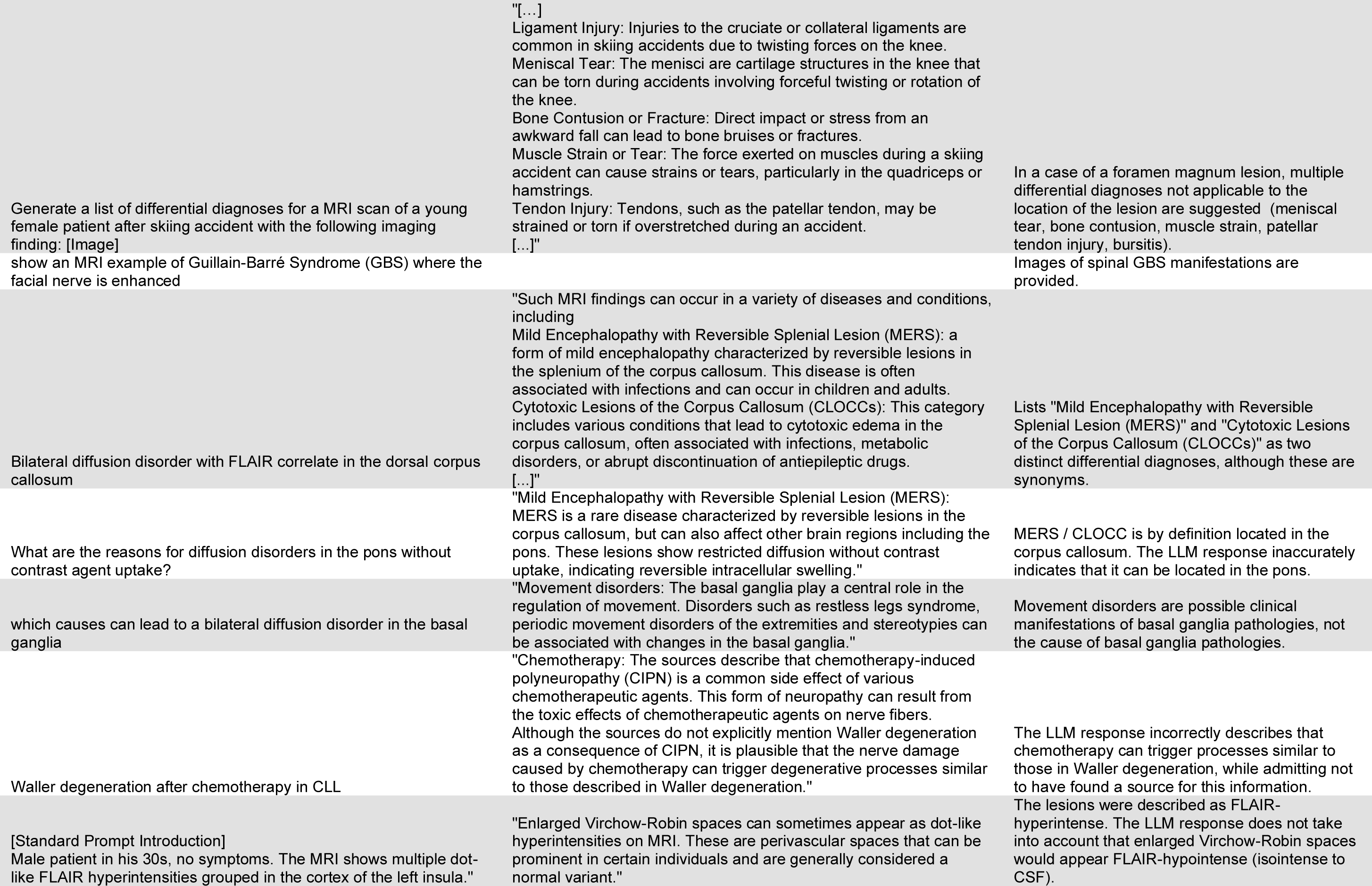
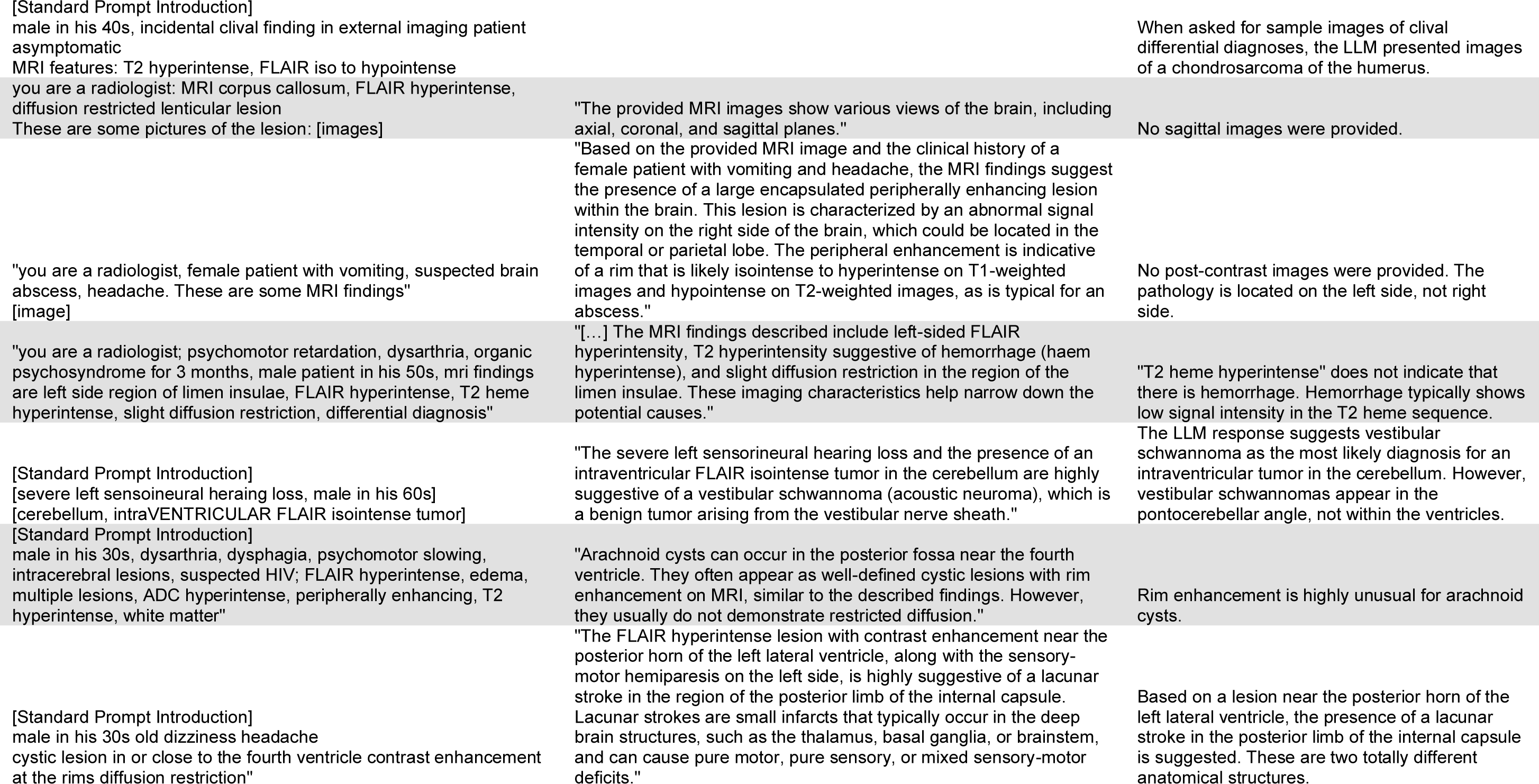
Hallucinatory LLM responses. “Standard prompt introduction” indicates the following part of the prompt template provided to the readers: “You are a senior neuroradiologist. Below, you will find information regarding a brain MRI scan. Based on this information, identify the three most likely differential diagnoses, ranked by their likelihood. Present your findings in a table format with the following columns: ‘Rank’, ‘Differential Diagnosis’, and ‘Explanation’.”

## References

1. Bhayana R. Chatbots and Large Language Models in Radiology: A Practical Primer for Clinical and Research Applications. Radiology. Radiological Society of North America Inc.; 2024;310(1). doi: 10.1148/RADIOL.232756/ASSET/IMAGES/LARGE/RADIOL.232756.FIG6.JPEG.

2. Gertz RJ, Bunck AC, Lennartz S, et al. GPT-4 for Automated Determination of Radiologic Study and Protocol Based on Radiology Request Forms: A Feasibility Study. Radiology. Radiological Society of North America Inc.; 2023;307(5). doi: 10.1148/RADIOL.230877.

3. Arachchige ASPM. Can We Use Large Language Models to Guide the Use of Contrast Media in Radiology? Reply to Kaba et al. Acad Radiol. Elsevier; 2024;0(0). doi: 10.1016/J.ACRA.2023.12.044.

4. Rau A, Rau S, Zöller D, et al. A Context-based Chatbot Surpasses Radiologists and Generic ChatGPT in Following the ACR Appropriateness Guidelines. Radiology. Radiological Society of North America Inc.; 2023;308(1). doi: 10.1148/RADIOL.230970.

5. Kumar Sarangi P, Irodi A, Panda S, Swapnesh Kumar Nayak D, Mondal H. Radiological Differential Diagnoses Based on Cardiovascular and Thoracic Imaging Patterns: Perspectives of Four Large Language Models. Indian Journal of Radiology and Imaging. 2023; doi: 10.1055/s-0043-1777289.

6. Horiuchi D, Tatekawa H, Shimono T, et al. Accuracy of ChatGPT generated diagnosis from patient’s medical history and imaging findings in neuroradiology cases. Neuroradiology. Neuroradiology; 2024;66(1). doi: 10.1007/S00234-023-03252-4.

7. Ueda D, Mitsuyama Y, Takita H, et al. ChatGPT’s Diagnostic Performance from Patient History and Imaging Findings on the Diagnosis Please Quizzes. Radiology. Radiological Society of North America Inc.; 2023;308(1). doi: 10.1148/RADIOL.231040.

8. Kottlors J, Bratke G, Rauen P, et al. Feasibility of Differential Diagnosis Based on Imaging Patterns Using a Large Language Model. Radiology. Radiological Society of North America Inc.; 2023;308(1). doi: 10.1148/radiol.231167.

9. Bhayana R, Bleakney RR, Krishna S. GPT-4 in Radiology: Improvements in Advanced Reasoning. Radiology. Radiological Society of North America Inc.; 2023;307(5). doi: 10.1148/RADIOL.230987.

10. Wu J, Kim Y, Keller EC, et al. Exploring Multimodal Large Language Models for Radiology Report Error-checking. 2023; https://arxiv.org/abs/2312.13103v1. Accessed January 14, 2024.

11. Ziegelmayer S, Marka AW, Lenhart N, et al. Evaluation of GPT-4’s Chest X-Ray Impression Generation: A Reader Study on Performance and Perception. J Med Internet Res. JMIR Publications Inc.; 2023;25(1):e50865. doi: 10.2196/50865.

12. Sun Z, Ong H, Kennedy P, et al. Evaluating GPT-4 on Impressions Generation in Radiology Reports. Radiology. Radiological Society of North America Inc.; 2023;307(5). doi: 10.1148/RADIOL.231259.

13. Huang J, Yang DM, Rong R, et al. A critical assessment of using ChatGPT for extracting structured data from clinical notes. npj Digital Medicine 2024 7:1. Nature Publishing Group; 2024;7(1):1–13. doi: 10.1038/s41746-024-01079-8.

14. Lehnen NC, Dorn F, Wiest IC, et al. Data Extraction from Free-Text Reports on Mechanical Thrombectomy in Acute Ischemic Stroke Using ChatGPT: A Retrospective Analysis. Anzai Y, editor. Radiology. Radiological Society of North America ; 2024;311(1). doi: 10.1148/RADIOL.232741.

15. Bhayana R, Elias G, Datta D, Bhambra N, Deng Y, Krishna S. Use of GPT-4 With Single-Shot Learning to Identify Incidental Findings in Radiology Reports. AJR Am J Roentgenol. American Roentgen Ray Society ; 2024; doi: 10.2214/AJR.23.30651/SUPPL_FILE/23_30651_SUPPL.PDF.

16. Wu J, Kim Y, Wu H. Hallucination Benchmark in Medical Visual Question Answering. 2024; https://www.openai.com/gpt-4. Accessed January 26, 2024.

17. Azamfirei R, Kudchadkar SR, Fackler J. Large language models and the perils of their hallucinations. Crit Care. BioMed Central Ltd; 2023;27(1):1–2. doi: 10.1186/S13054-023-04393-X.

18. Shen Y, Heacock L, Elias J, et al. ChatGPT and Other Large Language Models Are Double-edged Swords. Radiology. Radiological Society of North America Inc.; 2023;307(2). doi: 10.1148/RADIOL.230163.

19. Bhayana R, Biswas S, Cook TS, et al. From Bench to Bedside With Large Language Models: AJR Expert Panel Narrative Review. AJR Am J Roentgenol. AJR Am J Roentgenol; 2024; doi: 10.2214/AJR.24.30928.

20. Cascella M, Montomoli J, Bellini V, Bignami E. Evaluating the Feasibility of ChatGPT in Healthcare: An Analysis of Multiple Clinical and Research Scenarios. J Med Syst. Springer; 2023;47(1):1–5. doi: 10.1007/S10916-023-01925-4/TABLES/2.

21. Omiye JA, Gui H, Rezaei SJ, Zou J, Daneshjou R. Large Language Models in Medicine: The Potentials and Pitfalls : A Narrative Review. Ann Intern Med. Ann Intern Med; 2024;177(2):210–220. doi: 10.7326/M23-2772.

22. Clusmann J, Kolbinger FR, Muti HS, et al. The future landscape of large language models in medicine. Communications Medicine 2023 3:1. Nature Publishing Group; 2023;3(1):1–8. doi: 10.1038/s43856-023-00370-1.

23. Kim SH, Schramm S, Berberich C, et al. Human-AI Collaboration in Large Language Model-Assisted Brain MRI Differential Diagnosis: A Usability Study. medRxiv. Cold Spring Harbor Laboratory Press; 2024;2024.02.05.24302099. doi: 10.1101/2024.02.05.24302099.

24. Schramm S, Preis S, Metz M-C, et al. Impact of Multimodal Prompt Elements on Diagnostic Performance of GPT-4(V) in Challenging Brain MRI Cases. medRxiv. Cold Spring Harbor Laboratory Press; 2024;2024.03.05.24303767. doi: 10.1101/2024.03.05.24303767.

25. Siepmann R, Huppertz M, Rastkhiz A, et al. The virtual reference radiologist: comprehensive AI assistance for clinical image reading and interpretation. Eur Radiol. Springer Science and Business Media Deutschland GmbH; 2024;1–15. doi: 10.1007/S00330-024-10727-2/TABLES/3.

26. Yablonski J. of UX. “ O’Reilly Media, Inc.”; 2024.

27. Wu C, Lei J, Zheng Q, et al. Can GPT-4V(ision) Serve Medical Applications? Case Studies on GPT-4V for Multimodal Medical Diagnosis. 2023; https://arxiv.org/abs/2310.09909v3. Accessed February 22, 2024.

28. Horiuchi D, Tatekawa H, Oura T, et al. Comparison of the diagnostic accuracy among GPT-4 based ChatGPT, GPT-4V based ChatGPT, and radiologists in musculoskeletal radiology. medRxiv. Cold Spring Harbor Laboratory Press; 2023;2023.12.07.23299707. doi: 10.1101/2023.12.07.23299707.

29. Brin D, Sorin V, Barash Y, et al. Assessing GPT-4 Multimodal Performance in Radiological Image Analysis. medRxiv. Cold Spring Harbor Laboratory Press; 2023;2023.11.15.23298583. doi: 10.1101/2023.11.15.23298583.

30. Deng J, Heybati K, Shammas-Toma M. When vision meets reality: Exploring the clinical applicability of GPT-4 with vision. Clin Imaging. Elsevier; 2024;108:110101. doi: 10.1016/J.CLINIMAG.2024.110101.

31. Perplexity Accelerates Foundation Model Training by 40% with Amazon SageMaker HyperPod | Perplexity Case Study | AWS. https://aws.amazon.com/solutions/case-studies/perplexity-case-study/. Accessed July 1, 2024.

32. Paranjape K, Schinkel M, Panday RN, Car J, Nanayakkara P. Introducing Artificial Intelligence Training in Medical Education. JMIR Med Educ. JMIR Publications Inc.; 2019;5(2). doi: 10.2196/16048.

33. Ngo B, Nguyen D, Vansonnenberg E. The Cases for and against Artificial Intelligence in the Medical School Curriculum. Radiol Artif Intell. Radiological Society of North America; 2022;4(5). doi: 10.1148/RYAI.220074.

34. Li Q, Qin Y. AI in medical education: medical student perception, curriculum recommendations and design suggestions. BMC Med Educ. BioMed Central Ltd; 2023;23(1):1–8. doi: 10.1186/S12909-023-04700-8/FIGURES/2.

35. Pinto Dos Santos D, Hempel JM, Mildenberger P, Klöckner R, Persigehl T. Structured Reporting in Clinical Routine. RoFo Fortschritte auf dem Gebiet der Rontgenstrahlen und der Bildgebenden Verfahren. Georg Thieme Verlag; 2019;191(1):33–39. doi: 10.1055/a-0636-3851.

36. Kohli M, Alkasab T, Wang K, et al. Bending the Artificial Intelligence Curve for Radiology: Informatics Tools From ACR and RSNA. Journal of the American College of Radiology. Elsevier; 2019;16(10):1464–1470. doi: 10.1016/J.JACR.2019.06.009.

37. ESR. ESR paper on structured reporting in radiology—update 2023. Insights Imaging. Springer; 2023;14(1). doi: 10.1186/S13244-023-01560-0.

38. Kim SH, Mir-Bashiri S, Matthies P, Sommer W, Nörenberg D. Integration of structured reporting into the routine radiological workflow. Radiologe. Springer Medizin; 2021;61(11):1005–1013. doi: 10.1007/S00117-021-00917-0/FIGURES/8.

39. Dikici E, Bigelow M, Prevedello LM, White RD, Erdal BS. Integrating AI into radiology workflow: levels of research, production, and feedback maturity. Journal of Medical Imaging. Society of Photo-Optical Instrumentation Engineers; 2020;7(1):1. doi: 10.1117/1.JMI.7.1.016502.

